# Incidence, risk and severity of SARS-CoV-2 reinfections in children and adolescents: a population-level study between March 2020 and July 2022

**DOI:** 10.1101/2022.10.09.22280690

**Authors:** Snežana Medić, Cleo Anastassopoulou, Zagorka Lozanov-Crvenković, Nataša Dragnić, Vladimir Petrović, Mioljub Ristić, Tatjana Pustahija, Athanasios Tsakris, John P. A. Ioannidis

## Abstract

**IMPORTANCE:** During the COVID-19 pandemic children and adolescents were massively infected worldwide. In 2022 reinfections became increasingly common and they may continue to be a main feature of the endemic phase of SARS-CoV-2. It is important to understand the epidemiology and clinical impact of reinfections.

**OBJECTIVE:** To assess the incidence, risk, and severity of SARS-CoV-2 reinfection in children and adolescents.

**DESIGN, SETTING, AND PARTICIPANTS:** A population-level observational study was performed using surveillance data from the Autonomous Province of Vojvodina, Serbia between March 6, 2020 and April 30, 2022 with follow-up until July 31, 2022. The population-based sample consisted of 32 524 residents of Vojvodina <18 years of age with laboratory confirmed SARS-CoV-2 infection.

**EXPOSURES:** The surveillance database of the Institute for Public Health of Vojvodina was harnessed for epidemiological data of laboratory-confirmed SARS-CoV-2 infections.

**MAIN OUTCOMES AND MEASURES:** Incidence rates of documented SARS-CoV-2 reinfection per 1000 person-months. Estimated risk of documented reinfection ≥90 days after laboratory confirmation of primary infection. Reinfection severity and associated hospitalizations and deaths.

**RESULTS:** A total of 964 children (3.0%) experienced documented reinfection. The incidence rate of SARS-CoV-2 documented reinfections was 3.2 (CI 3.0-3.4) cases per 1000 person-months and was highest in adolescents aged 12-17 years (3.4; CI 3.2-3.7). Most reinfections (905, 93.9%) were recorded in 2022. The reinfection risk was 1.3% at six months, 1.9% at nine months, 4.0% at 12 months, 6.7% at 15 months, 7.2% at 18 months and 7.9% after 21 months.

Pediatric COVID-19 cases were generally mild. The proportion of severe clinical forms decreased from 14 (1.4%) in initial episodes to 3 (0.3%) in reinfections. Reinfected children were 4.7 times more likely to suffer from severe disease during initial infection compared to reinfection (McNemar OR=4.7; 95%CI 1.3-16.2, *p*=0.015). Pediatric reinfections rarely led to hospitalization (0.5% *vs*. 1.3% during primary infections) and none resulted in death.

**CONCLUSION AND RELEVANCE:** Reinfections are becoming more frequent as the pandemic progresses, yet the risk remains substantially lower for children and adolescents compared to adults. Pediatric infections rarely had clinical consequences and reinfections were even milder than primary infections.

**Key Points:** *Question:* What is the incidence, risk and severity of SARS-CoV-2 reinfection in children and adolescents?

*Findings:* This observational population-level study showed that the risk of pediatric reinfection remained less than 8% two years into the pandemic with an incidence rate of 3.2 (CI 3.0-3.4) cases per 1000 person-months. Pediatric COVID-19 cases were generally mild and reinfected children were 4.7 times more likely to suffer from severe disease during the initial infection compared to reinfection.

*Meaning:* These findings suggest that documented reinfection risk remains substantially lower in the pediatric versus the adult population, with an even more favorable profile compared to primary infections.

## Introduction

Age is an important parameter affecting the clinical outcome of severe acute respiratory syndrome coronavirus 2 (SARS-CoV-2) infections and coronavirus disease 2019 (COVID-19).^1^ In most children and adolescents, SARS-CoV-2 infections are mild or asymptomatic, although severe illness, including a rare multisystem inflammatory syndrome (MIS-C), as well as prolonged sequelae have been observed, particularly among young children.^2,3^ Despite the perhaps inevitable underreporting of subclinical infections, surveillance data from the pediatric population provided real-time information on symptomatic infections during the pandemic, informing risk assessments of the spread of the virus in schools.^4,5^

Data on SARS-CoV-2 reinfection in pediatric cohorts are scarce and mainly cover the pre-Omicron period. These studies showed a lower risk of reinfection for children compared to adults and no association with more severe disease or fatal outcomes.^6,7^ A surveillance study from England concluded that the rates of SARS-CoV-2 reinfection followed community infection rates.^6^ Symptomatic reinfections represented a large proportion of symptomatic COVID-19 cases (19.8%) among children in Nicaragua.^2^ Overall, the risk and severity of SARS-CoV-2 reinfection in children remain poorly characterized.

We aimed to assess the incidence, severity and risk of SARS-CoV-2 reinfection in children and adolescents by analyzing surveillance data for the Autonomous Province of Vojvodina, Serbia, from the beginning of the pandemic (March 6, 2020) to the end of July 2022.

## Methods

### Study design, objectives, and data collection

We conducted a retrospective population-level study to determine the incidence, risk and severity of SARS-CoV-2 reinfections in children and adolescents <18 years in Vojvodina, Serbia between March 6, 2020, and April 30, 2022. Reinfections were monitored until the end of follow-up (July 31, 2022) or date of death. Primary or first infection was defined as the detection of SARS-CoV-2 RNA or antigen in nasopharyngeal swab samples. SARS-CoV-2 reinfection was defined based on a positive RT-PCR or rapid antigen test (Ag-RDT) result ≥90 days after laboratory confirmation of primary infection.^6,8^

The study was conducted at the Institute for Public Health of Vojvodina (IPHV), an institution that oversights surveillance of infectious diseases among all ~1.9 million inhabitants of Vojvodina (comprising 26.7% of the population of Serbia).^9^ The surveillance IPHV database contains demographic and epidemiological data for all SARS-CoV-2 laboratory-confirmed Vojvodina residents from March 6, 2020. Data originated from epidemiological questionnaires and included information on patients’ gender, age, residence, description of symptoms, including date of symptom onset, illness severity, presence and number of comorbidities, hospitalization, date of SARS-CoV-2 laboratory confirmation, used diagnostic test, date of case registration, and disease outcome (recovery or death within 90 days of testing positive for SARS-CoV-2). The cause of death was classified as either COVID-19 or non-COVID, according to the information on the patients’ death certificates.

For surveillance purposes, the severity of COVID-19 was classified as: mild, if no pneumonia was diagnosed by chest imaging; severe, if COVID-19 pneumonia was confirmed by chest imaging; and critical, if COVID-19 illness required admission to the intensive care unit (ICU) and/or mechanical ventilation. A Unique Master Citizen Number (the official personal 13-digit code for Serbian citizens) was used to search the surveillance database for documented reinfections and to link recorded cases with the national registry of administered COVID-19 vaccines. Obtained data on vaccination status included the dates and type of vaccine administered. For 4.8% of cases for which the ID number was missing, the search was performed by patient name, surname, date of birth and residence. Overall, 1258 primary infections were excluded due to the limited time for reinfection to occur. The distribution of SARS-CoV-2 variants in Europe during pandemic waves is shown in eAppendix 1 in the Supplement.

### COVID-19 diagnostic algorithm

The COVID-19 diagnostic algorithm in Vojvodina has been previously described.^10^ In brief, laboratory-confirmed cases were defined based on SARS-CoV-2 positive RT-PCR or Ag-RDTs of posterior nasopharyngeal swab samples within the first five days of illness. Patients with COVID-19 symptoms and negative Ag-RDT results were tested by RT-PCR in repeated nasopharyngeal swabs.

### Classification of cases by vaccination status

In Serbia, vaccination against COVID-19 (with mRNA vaccines only) has been available since February 2021 for adolescents aged 16-17 years and since June 2021 for children ≥12 years, with a booster dose introduced in March 2022. Vaccination status (unvaccinated, partially or fully vaccinated and boosted) was defined according to the number of received vaccine doses in relation to the date of appearance of reinfection symptoms (or a positive test in the absence of symptoms).

### Statistical analysis

The share of documented primary SARS-CoV-2 infections and reinfections in the pediatric population was examined as proportions. Descriptive statistics of demographic characteristics of participants were conducted. The following pre-specified age groups were analyzed: 0-1, 2-11, 12-17 years. Examined variables included the presence of comorbidities and COVID-19 severity, including hospitalization. The trend of proportions of primary infection that were reinfected was assessed by the Cochran-Armitage test. Paired McNemar tests were used to compare reinfection versus primary infection in analyses with severe disease and hospitalization outcomes.

Incidence rates of reinfection were expressed per 1000 person-months. We also calculated age-specific incidence of primary infection and reinfection per 100 000 inhabitants using the 2021 population estimates as denominators.^9^ The average time-period between primary infection and reinfection was calculated. Time-to-event Kaplan-Meier analysis was used to estimate the reinfection risk during follow-up, starting at three months after primary infection. Using the same criteria, we also estimated the risk of severe/critical disease over time. COVID-19 vaccine uptake in the pediatric cohort was followed from mid-February 2021 until July 31, 2022.

Our final dataset was organized with Microsoft Access. A p-value <0.005 was considered statistically significant and p-values <0.05 were considered suggestive.^11^ Stata v.16 (College Station, TX: StataCorp LLC. 2019) and TIBCO Statistica™ 14.0.0 (license for University of Novi Sad) were used for statistical analyses.

### Ethics Statement

The collection of samples for COVID-19 diagnosis formed part of the standard patient management in the frame of national public health surveillance. Only oral informed consent was required and obtained from patients (or legal guardians of children aged <16). Patients’ data were anonymized before analysis. Data were stored in a secure server accessible only to authorized IPHV employees. Ethical approval for this study was waived by the Ethics Committee of IPHV.

## Results

### Primary SARS-CoV-2 infections and reinfections in the population of Vojvodina

A total of 456 334 COVID-19 cases were registered among the population of Vojvodina from March 6, 2020 until July 31, 2022 (eTable 1 in the Supplement). The proportion of SARS-CoV-2 reinfections was 26 016 (5.7%). Reinfections were rare in 2020 (25, 0.03%), increased about 30-fold in 2021 (1856, 0.9%), and almost 500-fold in 2022 (24 135, 14.7%); approximately half of all 2022 reinfections were recorded in January (11 967, 49.6%).

The cohort included a subset of 33 782 of 456 334 COVID-19 cases (7.4%) registered in children and adolescents <18 years (eTable 1). Increases were noted in 2021 in the recorded share of pediatric cases (from 2047 of 78 845 [2.6%] in 2020 to 18 967 of 213 645 [8.9%] in 2021), with surges In September and October (5015 and 6506 of 18 967 [26.4% and 34.3%], respectively) (eTable 1, **Figure 1A**). In 2022, pediatric cases remained at similar levels (12 768 of 163 844 [7.8%]) with most infections (7638 of 12 768 [59.8%]) recorded in January 2022.

**Figure 1.**
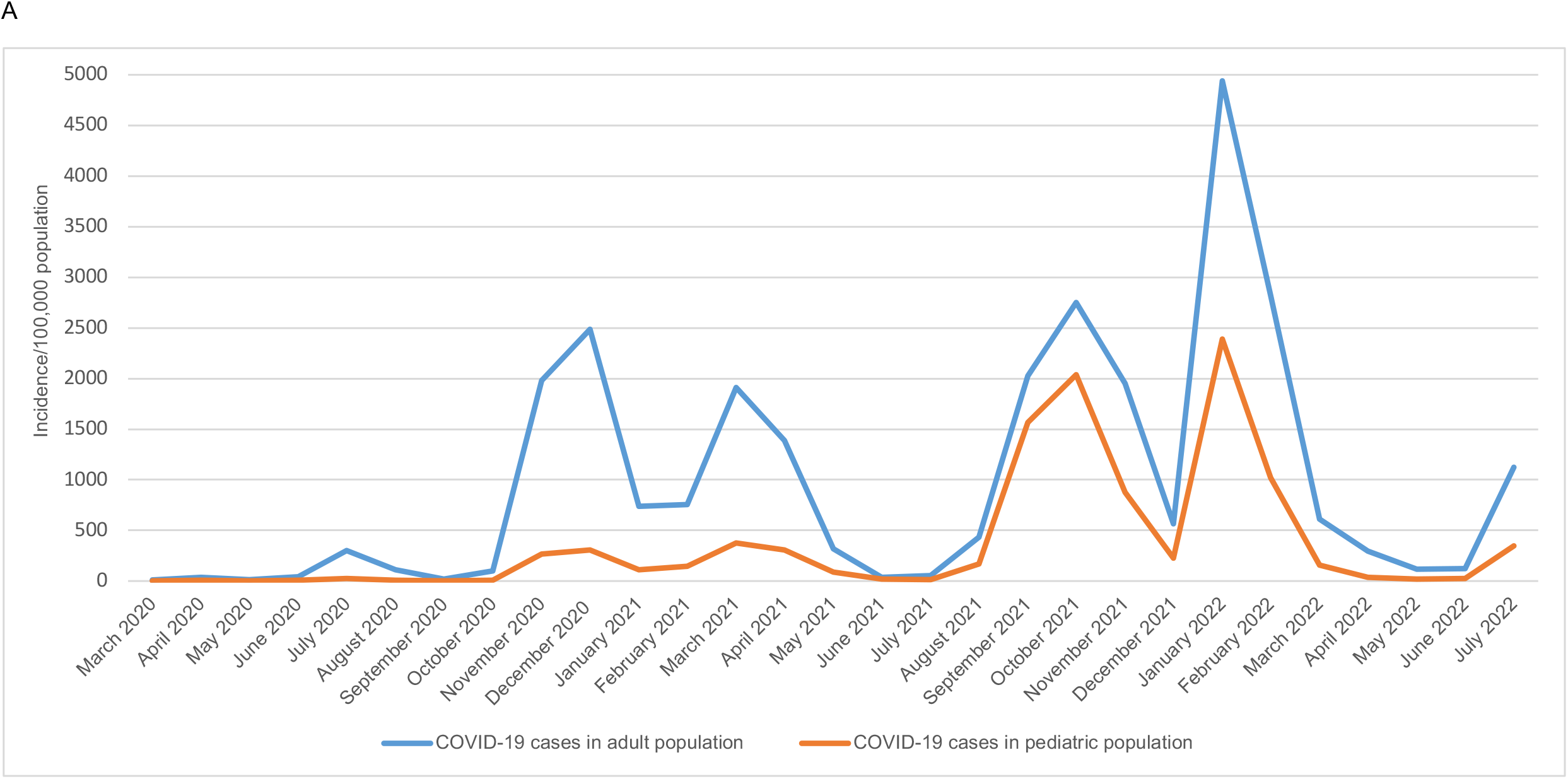
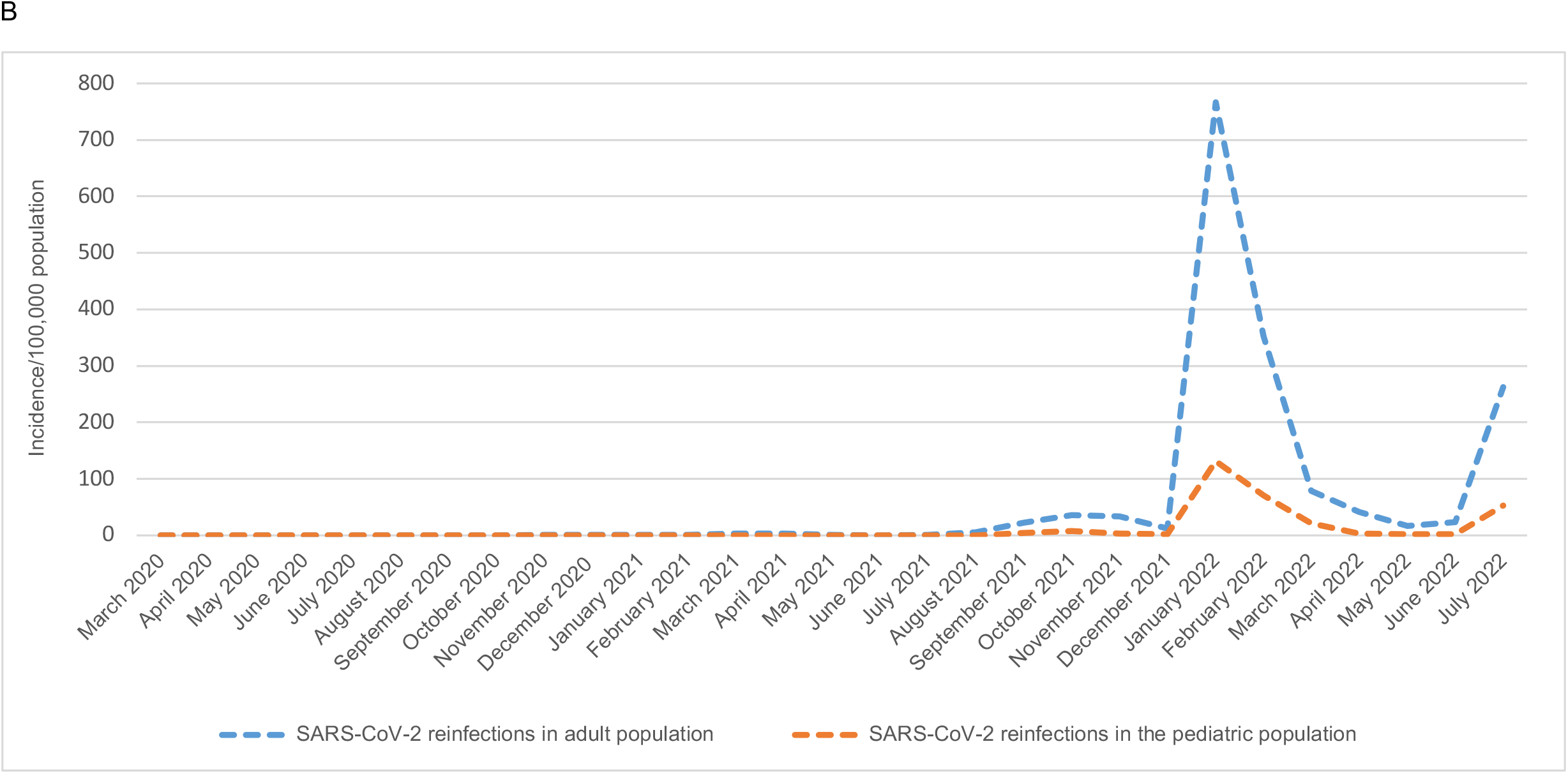
Incidence of (A) COVID-19 cases^a^ and (B) SARS-CoV-2 reinfections in the pediatric^b^ and adult^c^ population of Vojvodina, Serbia, March 6, 2020-July 31, 2022. ^a^ Excluding primary SARS-CoV-2 infections that ended in reinfections ^b^ Children and adolescents aged <18 years ^c^ Aged ≥18 years

Of the total of 32 524 primary SARS-CoV-2 infections registered among children and adolescents until April 30, 2022, 964 (3.0%) experienced reinfection (eTable 2). The average time duration from primary infection to reinfection was 240±117 days (min 90 days–max 591 days). Most infections were confirmed by Ag-RDTs (30 388 of 32 524 [93.4%]). No reinfections were documented in the pediatric cohort in 2020, very few (59 of 18 967 [6.1%]) in 2021, while most (905 of 964, [93.9%]) were recorded in 2022 (eTable 2). The highest proportion of primary infections that were reinfected was recorded in June 2021 (7 of 55 [12.7%]) during the fourth pandemic wave, with decreasing trends afterwards (eTable 2, eFigure 1).

### Incidence and estimated risk of SARS-CoV-2 reinfections in children and adolescents

Similar incidence patterns of primary infection and reinfection were noted in the pediatric versus the adult population, but to a lower extent in the former (**Figure 1B**). In adults, reinfections surged during the sixth (Jan. 1-Jun. 30, 2022) and seventh (July 2022-) pandemic waves, with peaks in January and July 2022 (766.6 and 262.1 per 100 000 inhabitants, respectively). This reinfection profile was mirrored in children and adolescents but at about 5.5-fold lower incidence. Pediatric reinfections also peaked in January 2022 (131.4/100 000 population). The highest incidence, recorded in 12-17-year-old adolescents, was 4.4-fold higher than the incidence in children aged 2-11 years (278.3 and 62.6/100 000 population, respectively). Both primary infection and reinfection rates were lower in younger (<12 years) pediatric age groups compared to older children (**Figure 2**).

**Figure 2.**
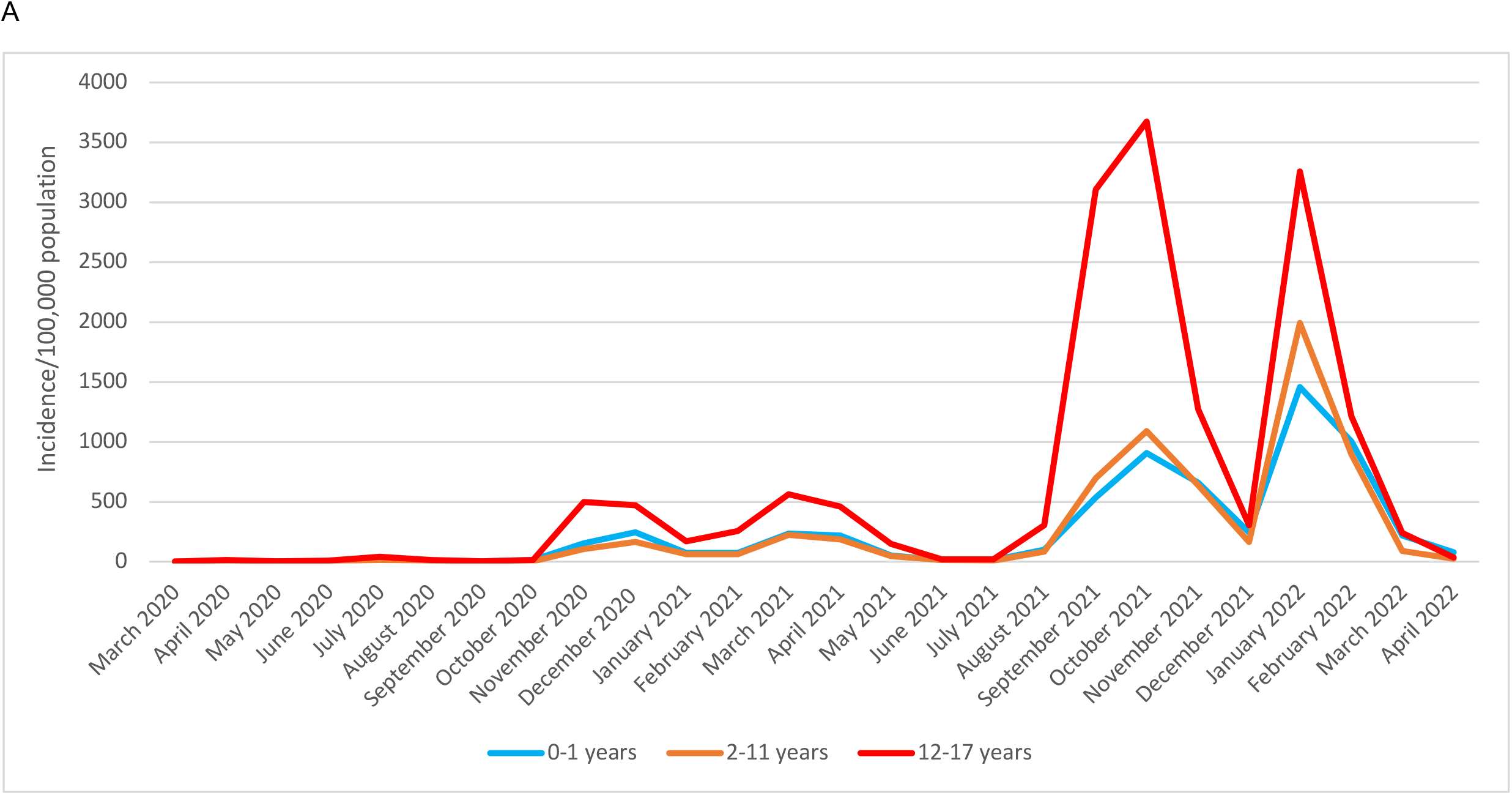
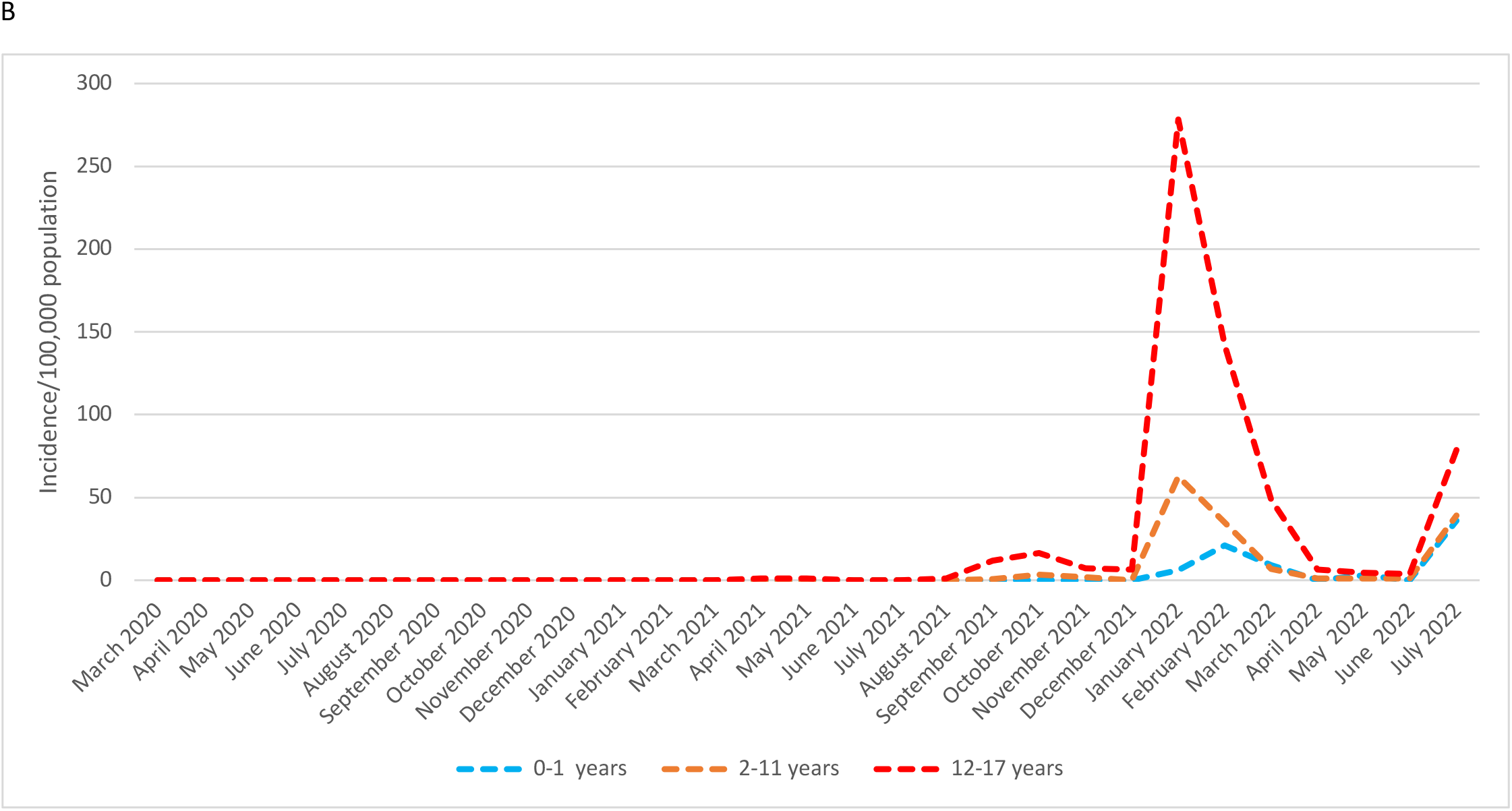
Age-specific incidences of (A) primary SARS-CoV-2 infections^a^ and (B) reinfections in children and adolescents, Vojvodina, Serbia, March 6, 2020-July 31, 2022. ^a^ Excluding primary SARS-CoV-2 infections that ended in reinfections.

The overall incidence rate of documented pediatric reinfections was 3.2 (CI 3.0-3.4) cases per 1000 person-months (**Table 1**). Rates did not differ much by gender, age, comorbidities, severity of primary infection or vaccination status. The highest rate was recorded in adolescents aged 12-17 years (3.4; CI 3.2-3.7). The mean age among participants with reinfections was 12.5±4.3 years (*vs*. 11.1±4.9 among cases without reinfection, *p*<0.001).

**Table 1.** Incidence Rates of SARS-CoV-2 Reinfection in Children and Adolescents in Vojvodina, Serbia, March 6, 2020-July 31, 2022.

| Characteristic | COVID-19 cases (N=32 524) | No. of reinfections (N=964) | No. of person-months (N=299 591) | Incidence rate of reinfections per 1000 person-months (95% CI) | Incidence rate ratio (95% CI) | p-value |
| --- | --- | --- | --- | --- | --- | --- |
| **Gender** |  |  |  |  |  |  |
| Male | 15 953 | 499 | 151 611 | 3.3 (3.0-3.6) | 1.1 (0.9-1.2) | 0.47 |
| Female | 16 571 | 465 | 147 980 | 3.1 (2.9-3.4) | Reference |  |
| **Age group (years)** |  |  |  |  |  |  |
| 0-1 | 2172 | 53 | 19 490 | 2.7 (2.0-3.6) | Reference |  |
| 2-11 | 11 920 | 305 | 103 483 | 2.9 (2.6-3.3) | 1.1 (0.8-1.5) | 0.60 |
| 12-17 | 18 432 | 606 | 176 618 | 3.4 (3.2-3.7) | 1.3 (0.9-1.7) | 0.097 |
| **Comorbidities<sup>a</sup>** |  |  |  |  |  |  |
| 0 | 31 818 | 936 | 291 903 | 3.2 (3.0-3.4) | Reference |  |
| ≥1 | 706 | 28 | 7688 | 3.8 (2.5-5.4) | 1.1 (0.8-1.7) | 0.50 |
| **Severity of disease<sup>b</sup>** |  |  |  |  |  |  |
| Mild | 32 182 | 950 | 293 629 | 3.2 (3.0-3.5) | 1.4 (0.8-2.5) | 0.063 |
| Severe | 342 | 14 | 5962 | 2.4 (1.3-3.9) | Reference |  |
| **Vaccination status<sup>c</sup>** |  |  |  |  |  |  |
| Unvaccinated | 32 315 | 963 | 298 197 | 3.2 (3.0-3.4) | 1.3 (0.2-52.9) | 0.88 |
| Partially vaccinated | 58 | 1 | 414 | 2.4 (0.0-13.5) | Reference |  |
| Fully vaccinated plus Boosted | 151 | 0 | 980 | 0 | 0 |  |

<sup>a</sup> Comorbidities included hypertension, diabetes, chronic pulmonary disease, chronic cardiovascular disease, malignancy, obesity, and other chronic diseases.
<sup>b</sup> There were no critical cases of COVID-19 in the pediatric population.
<sup>c</sup> At the time of laboratory confirmation of the primary infection, using the following definitions: Unvaccinated included those who had received 1 dose ≤14 days before the date of reinfection symptoms onset (or positive test in asymptomatic cases). Partially vaccinated: if 1 dose was received and >14 days had passed since the vaccination day; or the vaccination schedule had not been completed; or if the final dose was given ≤14 days or >6 months before the reinfection date. Fully vaccinated: if 2 vaccine doses were administered
and the final dose was received $>14$ days and $\leq 6$ months before the reinfection date. Boosted: if $>7$ days had passed since receiving the third (booster) dose before the reinfection date.

**Figure 3A** shows the Kaplan-Meier plot for the reinfection risk for pediatric patients who survived three months after primary infection (n=32 521). The risk becomes 1.3% at six months, 1.9% at nine months, 4.0% at 12 months, 6.7% at 15 months, 7.2% at 18 months and remains the same after 21 months (7.9%) to the end of follow up. The probability of reinfection was highest for children and adolescents who experienced primary infection during the third (Oct. 7, 2020-Jan. 31, 2021) and fourth pandemic waves (Feb. 1-Jul. 23, 2021) (**Figure 3B**). The risk of reinfection did not differ substantially for patients who suffered from mild versus severe primary infections (eFigure 2A), in different age groups (eFigure 2B), or between the sexes (eFigure 2C).

**Figure 3.**
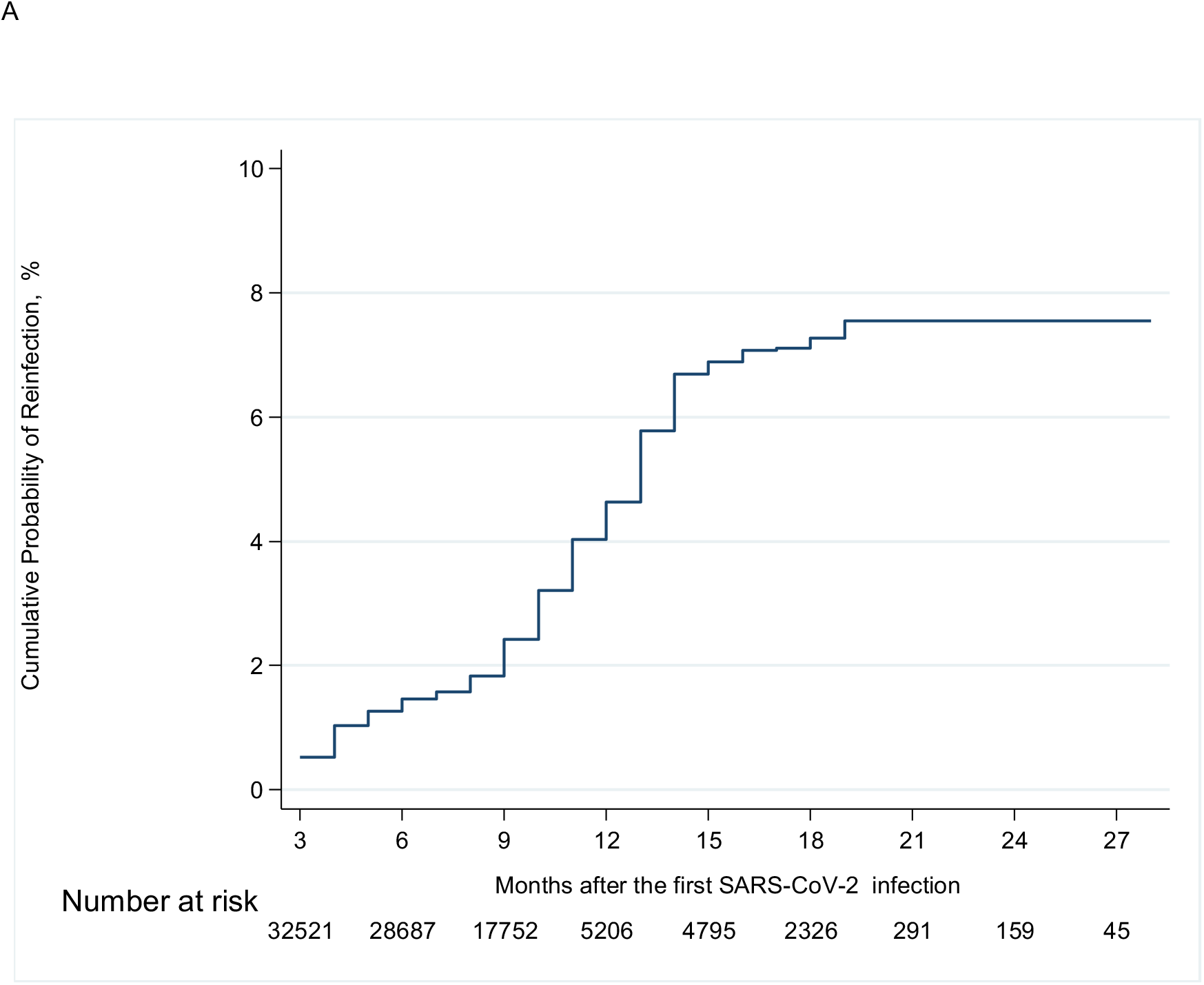
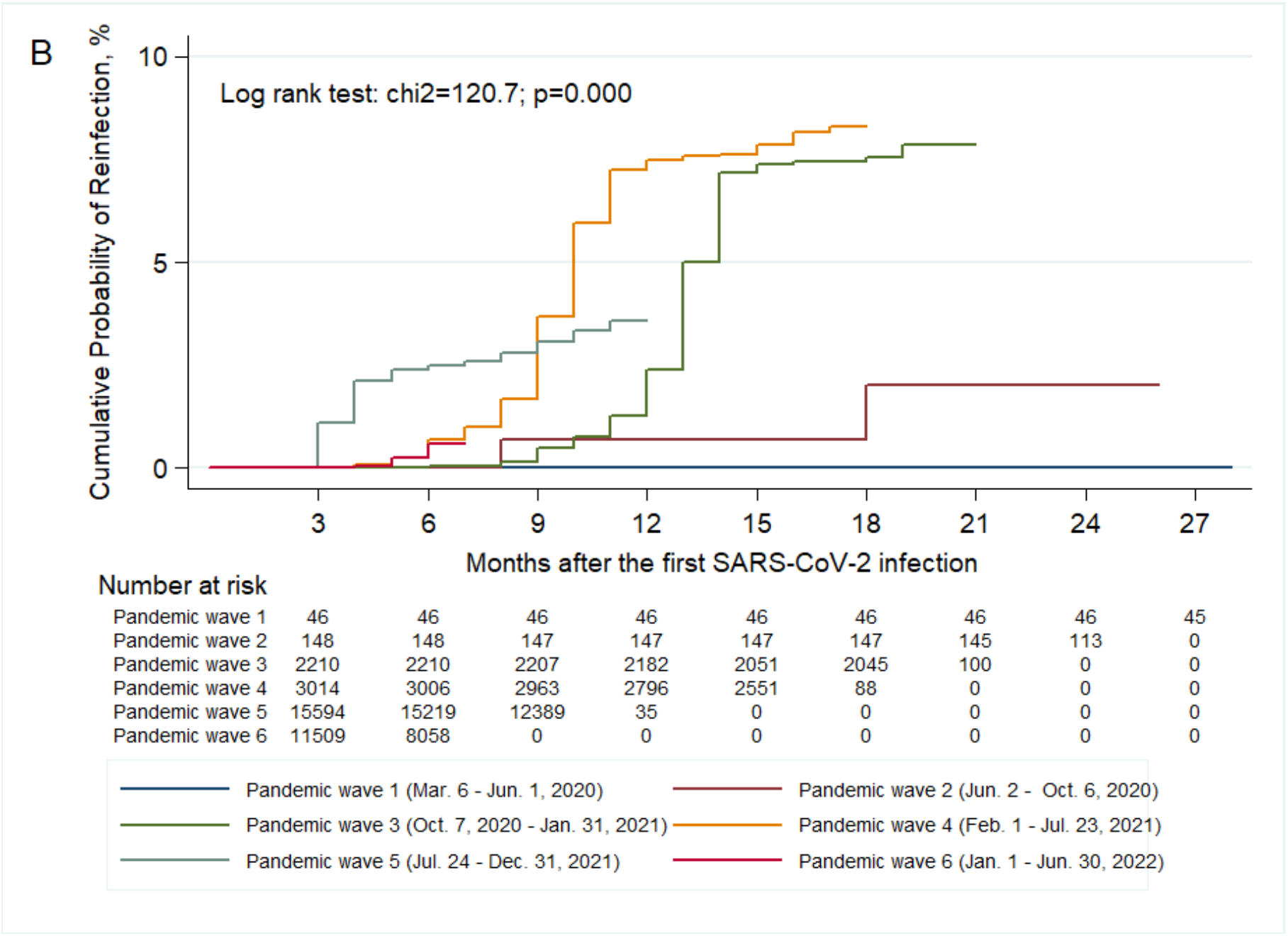
Kaplan-Meier Curves showing (A) the Cumulative Probability of Reinfection in the pediatric cohort (<18 years of age) and (B) According to Pandemic Waves in Vojvodina, Serbia, March 6, 2020-July 31, 2022.

### Severity of pediatric reinfections compared to primary infections

Pediatric COVID-19 cases were generally mild (**Table 1**, eFigure 3). The share of severe clinical forms decreased from 14 (1.4%) in initial episodes to 3 (0.3%) in reinfections. Reinfected patients were 4.7 times more likely to suffer from severe disease during initial infection compared to reinfection (McNemar OR=4.7; 95%CI 1.3-16.2, *p*=0.015).

### Hospitalizations after reinfection

There were 426 hospitalizations among 32 524 pediatric COVID-19 patients (with primary SARS-CoV-2 infection) in the observed period (1.3%) (eTable 2). At the beginning of the pandemic (Mar.-Jun. 2020), all SARS-CoV-2-positive children were hospitalized (due to official protocols that required hospitalization of COVID-19 cases in this early phase). The proportion of hospitalized patients decreased 10-fold, from 164 of 2047 (8.0%) in 2020 to 157 of 18 967 (0.8%) in 2021 and remain at a similar level in 2022 (105 of 11 510 [0.9%]). Reinfected children were rarely hospitalized: only five hospitalizations were recorded (0.5%), during the sixth and the seventh pandemic wave. An infant with comorbidities was hospitalized during both episodes. Kaplan-Meier curves for the risk of hospitalization for the entire cohort of 32 524 positive patients and for reinfected patients are shown in eFigure 4.

### Deaths

Three fatal outcomes were recorded among 32 524 SARS-CoV-2-positive children, of which one was classified as a COVID-19 death (case fatality ratio=0.003%). This child that suffered from a serious congenital heart defect died from pneumonia caused by COVID-19 in February 2022. The other two children died of non-COVID-19 causes. All three deaths were recorded during the sixth pandemic wave. No deaths occurred among reinfected children and adolescents.

### Second reinfections

Nine of 32 524 (0.03%) cases with three consecutive SARS-CoV-2 reinfections were recorded during the study (eTable 3). Cases of second reinfection ≥90 days after the previous episode were registered during the sixth (n=4) and seventh pandemic wave (n=5). Second reinfections were all mild and none led to hospitalization. On the day of laboratory confirmation of the second reinfection, most patients (8 of 9 [88.9%]) were unvaccinated.

## Discussion

Our population-level study showed that the risk of documented reinfection in children and adolescents was low before the advent of Omicron in January 2022. The risk of documented pediatric reinfection remained less than 8% two years into the pandemic, which is substantially lower than the risk of documented reinfection in the adult population. Both primary infections and reinfections in children and adolescents almost always had no clinical consequences. Severe infections and hospitalizations were rare and only one child died of COVID-19. Almost all severe infections and hospitalizations and the single death occurred upon primary infection. Reinfections had an even more favorable clinical course than primary infections.

A review of studies worldwide suggests that in the early pandemic phase (Jan.-Mar., 2020), children and adolescents accounted for less than 5% of documented COVID-19 cases. Increases were noted over time, and as of April 2022 children and adolescents accounted for 10-23% of total COVID-19 cases.^12^ This increase may largely reflect the wider use of testing with more frequent detection of asymptomatic cases, since the share of asymptomatic infections is larger in children than in adults.^13^ To some extent it may also reflect the opening of schools and greater socialization of children as the pandemic progressed.

Data on pediatric reinfections presented to-date covered mainly the pre-Omicron period. Comparisons should be cautious due to differences in methodology, including reinfection case definition, follow-up periods and limitations of community testing. In 2020-2021, SARS-CoV-2 pediatric reinfections were uncommon and with milder clinical course compared to primary infection.^7^ Mensah *et al*. have shown a higher incidence of documented reinfection in adults compared to children as well as an increase in reinfection rate with age.^6^ In another US-based series of pediatric reinfections until mid-2021, reinfections were not associated with severe or fatal outcomes.^14^ In our data, we observed an increase in the proportion of pediatric COVID-19 cases from 2.6% in 2020 to 8.9% in 2021. Testing in Serbia was limited across age groups in 2020 and became more widely available in 2021,^10^ covering a larger share of children. In 2022, pediatric COVID-19 infections in Vojvodina accounted for only 7.8%, which tends to be lower than estimates from other countries.^12^ The lower risk of reinfection in children and adolescents than in adults may reflect a more rigorous and more lasting immune response in younger populations. Alternatively, it may herald a larger share of missed (presumably asymptomatic) reinfections in children or adolescents than in adults. If so, this would strengthen even more the observation that pediatric reinfections have very benign clinical outlook.^1^

COVID-19 mitigating strategies, including school closures, were relaxed in Serbia in 2021. The number of symptomatic pediatric cases that would require testing increased substantially in 2021. In September/October 2021, when schools reopened, the share of symptomatic pediatric cases increased sharply (26.4% and 34.3% respectively), but the hospitalization rate was low (0.5% for primary infections and zero for reinfections). However, ~24% of all pediatric primary infections and ~44% of reinfections were registered in January 2022, following the emergence of Omicron, when most schools resumed in-person teaching. Even then, the hospitalization rate remained at the same low levels. The explosion of reinfections caused by Omicron and its subvariants stems from the evasion of both natural and vaccine-induced immunity, often within weeks after the previous episode, thus challenging the current >90 days reinfection case definition.^12,15–17^ Several studies indicated that children infected with Omicron are less likely to experience severe disease requiring hospitalization.^18–20^ This may reflect less aggressive features of the Omicron variant and/or natural immunity from undetected prior infecitons. Perhaps a considerable share of the Omicron infections that were coded as primary infections were in fact reinfections following undocumented primary infections in 2020-2021.

High rates of transmission of SARS-CoV-2 in the community were accompanied by outbreaks in schools. Children and adolescents have been probably almost ubiquitously infected by mid-2022 despite implemented mitigation strategies in schools.^21–23^ In the US, seroprevalence studies suggest that 75% of children and adolescents had antibodies by February 2022.^24^ Children and adolescents seem to have exhibited largely the same infection rates as adults during the pandemic^25^ and vaccinations may not have influenced this pattern. The vaccination rate in Serbia was stagnant in 2022 (47.8% completely vaccinated by June 2022),^26^ and vaccination coverage of children was negligible (<1%).

Some limitations of our work should be discussed. First, documentation of both primary infections and reinfections was limited by the availability of testing and unavoidably many subclinical infections were missed. Thus, the rates of clinically severe disease and complications are even lower than the extremely low rates that we observed. If the proportion of missed asymptomatic non-tested reinfections is higher in children than in adults, the lower rate of observed reinfections in children may be an artifact. Second, hospitalization rates depend on the threshold for admission. All children with COVID-19 were hospitalized in the first three months of the pandemic based on precautionary policy choice; hospitalization rates plummeted subsequently and were close to zero for reinfections. Comparison of hospitalization data for children with other countries is precarious because it is difficult to distinguish true differences in severity from differences in admission thresholds. For example, hospitalization rates for children and adolescents with COVID-19 have been much higher in the US,^12,27^ but many admissions might have been due to diseases other than COVID-19.^28^ Many recorded pediatric COVID-19 deaths may also be due to other diseases. This may explain the spuriously higher rates of recorded COVID-19 deaths in USA compared with European countries, including Serbia.^29^

Third, we have not captured information on potential long-term consequences of COVID-19 infection in children and adolescents, although claims for frequent pediatric long-COVID-19 may be unfounded.^30^ Fourth, children and adolescents in the studied population were mostly unvaccinated and our analyses were underpowered to address the impact of vaccination. However, outcomes were already excellent for pediatric COVID-19 infections and even more so for reinfections.

Acknowledging these caveats, our study maps pediatric COVID-19 infections and reinfections until July 2022 in a large population and offers reassuring evidence on the mild nature of reinfections. Further follow-up may offer insights into patterns of primary infections and reinfections, as new viral variants emerge.

## Supporting information

eAppendix 1, eTables 1-3, eFigures 1-4

## Data Availability

All data produced in the present study are available upon reasonable request to the authors

## Authors’ contributions

SM, CA and JPAI conceived and designed the study. SM, ND, VP, MR and TP processed laboratory results and collected data, SM, CA, ZLC, ND and AT analyzed and interpreted data, SM and CA supervised and validated data. SM, CA, ZLC, ND, TP and JPAI were involved in data visualization, presentation and formal analysis. SM, ZLC, ND and VP accessed and verified underlying data. SM, CA, AT and JPAI drafted the Article. All authors critically reviewed the article. All authors had access to aggregated data reported in the study and had final responsibility for the decision to submit the publication.

## Data sharing statement

The data that support the findings of this study are available upon request with approval needed from the Center for Disease Control and Prevention, Institute of Public Health of Vojvodina. The data are not publicly available due to restrictions pertaining to contained information that could compromise the privacy of patients.

## Acknowledgements

The authors would like to thank all epidemiologists and health-care workers who were involved in the study. The work of Zagorka Lozanov-Crvenković was funded by the Ministry of Education, Science and Technological Development of the Republic of Serbia (grant numbers: 451-03-68/2022-14/200032 and 451-03-68/2022-14/200125).

## References

1. Chou J, Thomas PG, Randolph AG. Immunology of SARS-CoV-2 infection in children. Nat Immunol. 2022; 23: 177–185. https://doi.org/10.1038/s41590-021-01123-9

2. Kubale J, Balmaseda A, Frutos AM, et al. Association of SARS-CoV-2 Seropositivity and Symptomatic Reinfection in Children in Nicaragua. JAMA Netw Open. 2022;5(6):e2218794. doi: 10.1001/jamanetworkopen.2022.18794

3. Anastassopoulou C, Spanakis N, Tsakris A. SARS-CoV-2 transmission, the ambiguous role of children and considerations for the reopening of schools in the fall. Future Microbiol. 2020;15:1201–1206. doi: 10.2217/fmb-2020-0195

4. Sah P, Fitzpatrick MC, Zimmer CF, et al. Asymptomatic SARS-CoV-2 infection: A systematic review and meta-analysis. Proc Natl Acad Sci U S A. 2021;118(34):e2109229118. doi: 10.1073/pnas.2109229118

5. Brodin P. SARS-CoV-2 infections in children: Understanding diverse outcomes. Immunity 2022;55(2):201–209. doi: 10.1016/j.immuni.2022.01.014

6. Mensah AA, Campbell H, Stowe J, et al. Risk of SARS-CoV-2 reinfections in children: a prospective national surveillance study between January, 2020, and July, 2021, in England. Lancet Child Adolesc Health. 2022;6(6):384–392. doi: 10.1016/S2352-4642(22)00059-1

7. Alhaddad F, Abdulkareem A, Alsharrah D, et al. Incidence of SARS-CoV-2 reinfection in a paediatric cohort in Kuwait. BMJ Open 2022;12:e056371. doi:10.1136/bmjopen-2021-056371

8. Yahav D, Yelin D, Eckerle I, et al. Definitions for coronavirus disease 2019 reinfection, relapse and PCR re-positivity. Clin Microbiol Infect. 2021;27(3):315–318.

9. Statistical Office of the Republic of Serbia. 2022. Estimates of population, 2021 [In Serbian/English]. Available from: https://publikacije.stat.gov.rs/G2022/PdfE/G20221172.pdf

10. Medić S, Anastassopoulou C, Lozanov-Crvenković Z et al. Risk and severity of SARS-CoV-2 reinfections during 2020-2022 in Vojvodina, Serbia: A population-level observational study. Lancet Reg Health Eur. 2022; 20:100453. doi: 10.1016/j.lanepe.2022.100453

11. Benjamin DJ, Berger JO, Johannesson M, et al. Redefine statistical significance. Nat Hum Behav. 2018;2(1):6–10.

12. Nathanielsz J, Toh ZQ, Do LAH, Mulholland K, Licciardi PV. SARS-CoV-2 infection in children and implications for vaccination. Pediatr Res. 2022;1–11. doi: 10.1038/s41390-022-02254-x

13. Yasuhara J, Kuno T, Takagi H, Sumitomo N. Clinical characteristics of COVID-19 in children: A systematic review. Pediatr Pulmonol. 2020;55(10):2565–2575. doi: 10.1002/ppul.24991

14. Comba IY, Riestra Guiance I, Corsini Campioli C, et al. Clinical Characteristics and Outcomes of Patients With SARS-CoV-2 Reinfection. Mayo Clin Proc Innov Qual Outcomes. 2022;6(4):361–372. doi: 10.1016/j.mayocpiqo.2022.05.004

15. Al-Otaiby M, Krissaane I, Al Seraihi A, et al. SARS-CoV-2 Reinfection Rate and Outcomes in Saudi Arabia: A National Retrospective Study. Int J Infect Dis. 2022;122:758–766. doi: 10.1016/j.ijid.2022.07.025

16. Chaguza C, Coppi A, Earnest R, et al. Rapid emergence of SARS-CoV-2 Omicron variant is associated with an infection advantage over Delta in vaccinated persons. Med (N Y). 2022;3(5):325–334.e4. doi: 10.1016/j.medj.2022.03.010

17. Pulliam JRC, van Schalkwyk C, Govender N, et al. Increased risk of SARS-CoV-2 reinfection associated with emergence of Omicron in South Africa. Science 2022;376(6593):eabn4947. doi: 10.1126/science.abn4947

18. Martin B, DeWitt PE, Russell S, et al. Acute Upper Airway Disease in Children With the Omicron (B.1.1.529) Variant of SARS-CoV-2-A Report From the US National COVID Cohort Collaborative. JAMA Pediatr. 2022;176(8):819–821. doi: 10.1001/jamapediatrics.2022.1110

19. Butt AA, Dargham SR, Loka S, et al. Coronavirus Disease 2019 Disease Severity in Children Infected With the Omicron Variant. Clin Infect Dis. 2022;75(1):e361–e367. doi: 10.1093/cid/ciac275

20. Wang L, Berger NA, Kaelber DC, Davis PB, Volkow ND, Xu R. Incidence Rates and Clinical Outcomes of SARS-CoV-2 Infection With the Omicron and Delta Variants in Children Younger Than 5 Years in the US. JAMA Pediatr. 2022;176(8):811–813. doi: 10.1001/jamapediatrics.2022.0945

21. Haapanen M, Renko M, Artama M, Kuitunen I. The impact of the lockdown and the reopening of schools and day cares on the epidemiology of SARS-CoV-2 and other respiratory infections in children - A nationwide register study in Finland. EClinicalMedicine 2021;34:100807. doi: 10.1016/j.eclinm.2021.100807

22. Macartney K, Quinn HE, Pillsbury AJ, et al. NSW COVID-19 Schools Study Team. Transmission of SARS-CoV-2 in Australian educational settings: a prospective cohort study. Lancet Child Adolesc Health 2020;4(11):807–816. doi: 10.1016/S2352-4642(20)30251-0

23. European Centre for Disease Prevention and Control (ECDC). COVID-19 in Children and the Role of School Settings in Transmission-Second Update (ECDC, Stockholm, 2021).

24. Clarke KEN, Jones JM, Deng Y, et al. Seroprevalence of Infection-Induced SARS-CoV-2 Antibodies - United States, September 2021-February 2022. MMWR Morb Mortal Wkly Rep. 2022;71(17):606–608. doi: 10.15585/mmwr.mm7117e3

25. Axfors C, Pezzullo AM, Contopoulos-Ioannidis DG, Apostolatos A, Ioannidis JPA. Differential COVID-19 infection rates in children, adults, and elderly: evidence from 38 prevaccination national seroprevalence studies. medRxiv 2022.06.28.22277034. doi: https://doi.org/10.1101/2022.06.28.22277034

26. Ritchie H, Ortiz-Ospina E, Beltekian D, et al. Coronavirus Pandemic (COVID-19); 2022. Available at: https://ourworldindata.org/coronavirus/country/serbia#what-share-of-the-population-has-completed-the-initial-vaccination-protocol (accessed September 27, 2022).

27. Marks KJ, Whitaker M, Anglin O, et al. Hospitalizations of Children and Adolescents with Laboratory-Confirmed COVID-19 - COVID-NET, 14 States, July 2021-January 2022. MMWR Morb Mortal Wkly Rep. 2022;71(7):271–278. doi: 10.15585/mmwr.mm7107e4

28. Klann JG, Strasser ZH, Hutch MR, et al. Distinguishing Admissions Specifically for COVID-19 From Incidental SARS-CoV-2 Admissions: National Retrospective Electronic Health Record Study. J Med Internet Res. 2022;24(5):e37931. doi: 10.2196/37931

29. Kitano T, Kitano M, Krueger C, et al. The differential impact of pediatric COVID-19 between high-income countries and low- and middle-income countries: A systematic review of fatality and ICU admission in children worldwide. PLoS One 2021;16(1):e0246326. doi: 10.1371/journal.pone.0246326

30. Hirt J, Janiaud P, Gloy VL, et al. Robustness of reported postacute health outcomes in children with SARS-CoV-2 infection: a systematic review. Archives of Disease in Childhood Published Online First: 02 September 2022. doi: 10.1136/archdischild-2022-324455

