## Supplementary material for "Incidence, risk and severity of SARS-CoV-2 reinfections in children and adolescents: a population-level study between March 2020 and July 2022": eAppendix 1, eTables 1-3, eFigures 1-4: Supplement_10.10.22.pdf

**eAppendix 1.** The Distribution of SARS-CoV-2 Variants in Europe during the Seven Pandemic Waves recorded in Serbia, March 6, 2020-January 31, 2022.
**eTable 1.** Primary SARS-CoV-2 Infections and Reinfections in Vojvodina, Serbia, March 6, 2020-July 31, 2022.
**eTable 2.** SARS-CoV-2 Primary Infections and Reinfections and Hospitalization Rates in the Pediatric Population of Vojvodina, Serbia, March 6, 2020-July 31, 2022.
**eTable 3.** Characteristics of Children and Adolescents with Two Consecutive SARS-CoV-2 Reinfections in Vojvodina, Serbia, March 6, 2020-July 31, 2022.
**eFigure 1.** The Proportion of Primary Infections that were Reinfected in Children, Vojvodina, Serbia, March 6, 2020-July 31, 2022.
**eFigure 2.** Kaplan-Meier Curves showing the Cumulative Probability of Reinfection in the pediatric cohort (<18 years of age) According to (A) Severity of Primary Infection, (B) Age Group and (C) Gender in Vojvodina, Serbia, March 6, 2020-July 31, 2022.
**eFigure 3.** Severity of COVID-19 in Children and Adolescents (<18 years) without and with SARS-CoV-2 Reinfection (first and second episode), in Vojvodina, Serbia, March 6, 2020-July 31, 2022.
**eFigure 4.** Kaplan-Meier Curves Showing the Cumulative Probability of Hospitalization(A) in the Pediatric Cohort and (B) for Patients with Reinfection in Vojvodina, Serbia, March 6, 2020-July 31, 2022.

**eAppendix 1. The distribution of SARS-CoV-2 variants in Europe during the Seven Pandemic Waves recorded in Serbia, March 6, 2020-January 31, 2022.** (Nextstrain naming was used).

1. First pandemic wave (Mar. 6 - Jun. 1, 2020): 19A, 19B, 20A, 20B, 20C, 20D.
2. Second pandemic wave (Jun. 2 - Oct. 6, 2020): 19B, 20A, 20B, 20C, 20D, 20E (EU1).
3. Third pandemic wave (Oct. 7, 2020 - Jan. 31, 2021): 19B, 20A, 20B, 20C, 20D, 20E (EU1), 20H (Beta, V2), 20I (Alpha, V1), 21C (Epsilon), 21D (Eta).
4. Fourth pandemic wave (Feb. 1 - Jul. 23, 2021): 19B, 20A, 20B, 20C, 20D, 20E (EU1), 20H (Beta, V2), 20I (Alpha, V1), 20J (Gamma, V3), 21A (Delta), 21I (Delta), 21J (Delta), 21B (Kappa), 21D (Eta), 21G (Lambda).
5. Fifth pandemic wave (Jul. 24 - Dec. 31, 2021): 19B, 20A, 20B, 20C, 20E (EU1), 20I (Alpha, V1), 20J (Gamma, V3), 21A (Delta), 21I (Delta), 21J (Delta), 21H (Mu), 21K (Omicron), 21L (Omicron).
6. Sixth pandemic wave (Jan. 1 - Jun. 30, 2022): 21I (Delta), 21J (Delta), 21K (Omicron), 21L (Omicron).
7. Seventh (ongoing) pandemic wave (Jul. 1, 2022 - ): 21L (Omicron), 22A (Omicron), 22B (Omicron).

**eTable 1. Primary SARS-CoV-2 Infections and Reinfections in Vojvodina, Serbia, March 6, 2020-July 31, 2022.**

| | Months | Overall COVID-19 cases[a] | | SARS-CoV-2 reinfections[b] | | Pediatric COVID-19 cases[a,c] | | Reinfections in the pediatric population[b,c] | | Share of pediatric cases in the total number of COVID-19 cases | Share of SARS-CoV-2 reinfections in children in the total number of reinfections |
|---|---|---|---|---|---|---|---|---|---|---|---|
| | | n | % | n | % | n | % | n | % | % | % |
| 2020 | March-June | 1570 | 0.4 | 0 | 0 | 68 | 0.2 | 0 | 0 | 4.3 | 0 |
| | July | 4597 | 1.0 | 0 | 0 | 83 | 0.3 | 0 | 0 | 1.8 | 0 |
| | August | 1750 | 0.4 | 0 | 0 | 33 | 0.1 | 0 | 0 | 1.9 | 0 |
| | September | 330 | 0.1 | 0 | 0 | 8 | 0.02 | 0 | 0 | 2.4 | 0 |
| | October | 1547 | 0.3 | 1 | 0.06 | 29 | 0.1 | 0 | 0 | 1.9 | 0 |
| | November | 30 659 | 6.7 | 9 | 0.03 | 854 | 2.5 | 0 | 0 | 2.8 | 0 |
| | December | 38 392 | 8.4 | 15 | 0.04 | 972 | 2.9 | 0 | 0 | 2.5 | 0 |
| | **2020 subtotal** | **78 845** | **17.3** | **25** | **0.03** | **2047** | **6.1** | **0** | **0** | **2.6** | **0** |
| 2021 | January | 11 441 | 2.5 | 7 | 0.06 | 357 | 1.1 | 0 | 0 | 3.1 | 0 |
| | February | 11 868 | 2.6 | 11 | 0.09 | 459 | 1.4 | 0 | 0 | 3.9 | 0 |
| | March | 30 012 | 6.6 | 47 | 0.2 | 1201 | 3.6 | 0 | 0 | 4.0 | 0 |
| | April | 21 929 | 4.8 | 47 | 0.2 | 983 | 2.9 | 1 | 0.1 | 4.5 | 2.1 |
| | May | 5084 | 1.1 | 21 | 0.4 | 285 | 0.8 | 1 | 0.4 | 5.6 | 4.8 |
| | June | 586 | 0.1 | 5 | 0.9 | 55 | 0.2 | 0 | 0 | 9.4 | 0 |
| | July | 860 | 0.2 | 8 | 0.9 | 47 | 0.1 | 0 | 0 | 5.5 | 0 |
| | August | 7029 | 1.5 | 80 | 1.1 | 544 | 1.6 | 1 | 0.2 | 7.7 | 1.3 |
| | September | 35 515 | 7.8 | 351 | 1.0 | 5015 | 14.8 | 14 | 0.3 | 14.1 | 4.0 |
| | October | 47 935 | 10.5 | 567 | 1.2 | 6506 | 19.3 | 24 | 0.4 | 13.6 | 4.2 |

| | Months | Overall COVID-19 cases[a] | | SARS-CoV-2 reinfections[b] | | Pediatric COVID-19 cases[a,c] | | Reinfections in the pediatric population[b,c] | | Share of pediatric cases in the total number of COVID-19 cases | Share of SARS-CoV-2 reinfections in children in the total number of reinfections |
|---|---|---|---|---|---|---|---|---|---|---|---|
| | | n | % | n | % | n | % | n | % | % | % |
| | **November** | 32 174 | 7.1 | 526 | 1.6 | 2798 | 8.3 | 11 | 0.4 | 8.7 | 2.1 |
| | **December** | 9212 | 2.0 | 186 | 2.0 | 717 | 2.1 | 7 | 1.0 | 7.8 | 3.8 |
| | **2021 subtotal** | **213 645** | **46.8** | **1856** | **0.9** | **18 967** | **56.1** | **59** | **0.3** | **8.9** | **3.2** |
| **2022** | **January** | 82 046 | 18.0 | 11 967 | 14.6 | 7638 | 22.6 | 420 | 5.5 | 9.3 | 3.5 |
| | **February** | 45 713 | 10.0 | 5535 | 12.1 | 3261 | 9.6 | 225 | 6.9 | 7.1 | 4.1 |
| | **March** | 9710 | 2.1 | 1257 | 12.9 | 500 | 1.5 | 69 | 13.8 | 5.2 | 5.5 |
| | **April** | 4562 | 1.0 | 639 | 14.0 | 111 | 0.3 | 9 | 8.1 | 2.4 | 1.4 |
| | **May** | 1829 | 0.4 | 263 | 14.4 | 70 | 0.2 | 8 | 11.4 | 3.8 | 3.0 |
| | **June** | 1924 | 0.4 | 358 | 18.6 | 76 | 0.2 | 6 | 7.9 | 3.9 | 1.7 |
| | **July** | 18 060 | 4.0 | 4116 | 22.8 | 1112 | 3.3 | 168 | 15.1 | 6.2 | 4.1 |
| | **2022 subtotal** | **163 844** | **35.9** | **24 135** | **14.7** | **12 768** | **37.8** | **905** | **7.1** | **7.8** | **3.8** |
| **Total 2020-2022** | | **456 334** | **100** | **26 016** | **5.7** | **33 782** | **100** | **964** | **2.9** | **7.4** | **3.7** |

[a] By month of first episode registration.
[b] By month of reinfection registration.
[c] Patients aged <18 years.

**eTable 2. SARS-CoV-2 Primary Infections and Reinfections and Hospitalization Rates in the Pediatric Population[a] of Vojvodina, Serbia, March 6, 2020-July 31, 2022.**

| | | Pediatric SARS-CoV-2 primary infections[b] | | Hospitalized primary infections[b, c] | | Proportion of SARS-CoV-2 reinfections in the total number of reinfections | | Proportion of SARS-CoV-2 reinfections in the number of primary infections[d] | | Proportion of hospitalized reinfections[c,d] | | Primary SARS-CoV-2 infections that were reinfected | |
|---|---|---|---|---|---|---|---|---|---|---|---|---|---|
| | **Months** | **n** | **%** | **n** | **%** | **n** | **%** | **n** | **%** | **n** | **%** | **n** | **%** |
| **2020** | March-June | 68 | 0.2 | 68 | 100.0 | 0 | 0 | 0 | 0 | 0 | 0 | 0 | 0 |
| | July | 83 | 0.3 | 45 | 54.2 | 0 | 0 | 0 | 0 | 0 | 0 | 2 | 2.4 |
| | August | 33 | 0.1 | 6 | 18.2 | 0 | 0 | 0 | 0 | 0 | 0 | 1 | 3.0 |
| | September | 8 | 0.02 | 1 | 12.5 | 0 | 0 | 0 | 0 | 0 | 0 | 0 | 0 |
| | October | 29 | 0.1 | 1 | 3.5 | 0 | 0 | 0 | 0 | 0 | 0 | 0 | 0 |
| | November | 854 | 2.6 | 21 | 2.5 | 0 | 0 | 0 | 0 | 0 | 0 | 65 | 7.6 |
| | December | 972 | 3.0 | 22 | 2.3 | 0 | 0 | 0 | 0 | 0 | 0 | 75 | 7.7 |
| | **Subtotal** | **2047** | **6.3** | **164** | **8.0** | **0** | **0** | **0** | **0** | **0** | **0** | **143** | **7.0** |
| **2021** | January | 357 | 1.1 | 4 | 1.1 | 0 | 0 | 0 | 0 | 0 | 0 | 33 | 9.2 |
| | February | 459 | 1.4 | 5 | 1.1 | 0 | 0 | 0 | 0 | 0 | 0 | 35 | 7.6 |
| | March | 1201 | 3.7 | 19 | 1.6 | 0 | 0 | 0 | 0 | 0 | 0 | 100 | 8.3 |
| | April | 983 | 3.0 | 17 | 1.7 | 1 | 0.1 | 1 | 0.1 | 0 | 0 | 79 | 8.0 |
| | May | 285 | 0.9 | 5 | 1.8 | 1 | 0.1 | 1 | 0.4 | 0 | 0 | 19 | 6.7 |
| | June | 55 | 0.2 | 3 | 5.5 | 0 | 0 | 0 | 0 | 0 | 0 | 7 | 12.7 |
| | July | 47 | 0.2 | 3 | 6.4 | 0 | 0 | 0 | 0 | 0 | 0 | 3 | 6.4 |
| | August | 544 | 1.7 | 6 | 1.1 | 1 | 0.1 | 1 | 0.2 | 0 | 0 | 23 | 4.2 |
| | September | 5015 | 15.4 | 24 | 0.5 | 14 | 1.5 | 14 | 0.3 | 0 | 0 | 175 | 3.5 |
| | October | 6506 | 20.0 | 31 | 0.5 | 24 | 2.5 | 24 | 0.4 | 0 | 0 | 227 | 3.5 |
| | November | 2798 | 8.6 | 27 | 1.0 | 11 | 1.2 | 11 | 0.4 | 0 | 0 | 53 | 1.9 |
| | December | 717 | 2.2 | 13 | 1.8 | 7 | 0.7 | 7 | 1.0 | 0 | 0 | 12 | 1.7 |
| | **Subtotal** | **18 967** | **58.3** | **157** | **0.8** | **59** | **6.1** | **59** | **0.3** | 0 | **0** | **766** | **4.0** |
| **2022** | January | 7638 | 23.5 | 37 | 0.5 | 420 | 43.6 | 420 | 5.5 | 4 | 1.0 | 42 | 0.6 |
| | February | 3261 | 10.0 | 46 | 1.4 | 225 | 23.3 | 225 | 6.9 | 0 | 0 | 11 | 0.3 |

| | | Pediatric SARS-CoV-2 primary infections[b] | | Hospitalized primary infections[b, c] | | Proportion of SARS-CoV-2 reinfections in the total number of reinfections | | Proportion of SARS-CoV-2 reinfections in the number of primary infections[d] | | Proportion of hospitalized reinfections[c,d] | | Primary SARS-CoV-2 infections that were reinfected | |
|---|---|---|---|---|---|---|---|---|---|---|---|---|---|
| | **Months** | **n** | **%** | **n** | **%** | **n** | **%** | **n** | **%** | **n** | **%** | **n** | **%** |
| | March | 500 | 1.5 | 16 | 3.2 | 69 | 7.2 | 69 | 13.8 | 0 | 0 | 2 | 0.4 |
| | April | 111 | 0.3 | 6 | 5.4 | 9 | 0.9 | 9 | 8.1 | 0 | 0 | 0 | 0 |
| | May | - | - | - | - | 8 | 0.8 | 8 | - | 0 | 0 | - | - |
| | June | - | - | - | - | 6 | 0.6 | 6 | - | 0 | 0 | - | - |
| | July | - | - | - | - | 168 | 17.4 | 168 | - | 1 | 0.6 | - | - |
| | **Subtotal** | **11 510** | **35.4** | **105** | **0.9** | **905** | **93.9** | **905** | **7.9** | **5** | **0.6** | **55** | **0.5** |
| **TOTAL** | | **32 524** | **100** | **426** | **1.3** | **964** | **100** | **964** | **3.0** | **5** | **0.5** | **964** | **3.0** |

[a] Aged <18 years.
[b] By month of first episode registration.
[c] Hospitalization within one month from the date of laboratory confirmation of infection or reinfection.
[d] By month of reinfection registration.

### eTable 3. Characteristics of Children and Adolescents[a] with Two Consecutive SARS-CoV-2 Reinfections in Vojvodina, Serbia, March 6, 2020-July 31, 2022.

| Gender | Age group (years)[b] | Number of comorbidities[b] | Vaccination status[b] | Time between infections (days) | | Pandemic wave[d] | | |
|---|---|---|---|---|---|---|---|---|
| | | | | 1st infection to 1st reinfection | 1st reinfection to 2nd reinfection | Primary infection[c] | 1st reinfection[c] | 2nd reinfection[c] |
| Male | 2-11 | 0 | Unvaccinated | 224 | 163 | 4 | 5 | 7 |
| Male | 2-11 | 0 | Unvaccinated | 131 | 129 | 5 | 6 | 6 |
| Male | 12-17 | 0 | Unvaccinated | 125 | 134 | 5 | 6 | 6 |
| Male | 12-17 | 1 | Unvaccinated | 494 | 132 | 3 | 6 | 7 |
| Female | 2-11 | 0 | Unvaccinated | 167 | 188 | 5 | 6 | 7 |
| Female | 12-17 | 0 | Unvaccinated | 121 | 135 | 5 | 6 | 7 |
| Female | 12-17 | 0 | Unvaccinated | 308 | 120 | 3 | 5 | 6 |
| Female | 12-17 | 0 | Unvaccinated | 149 | 164 | 5 | 6 | 7 |
| Female | 12-17 | 0 | Partially vaccinated[e] | 208 | 319 | 4 | 5 | 6 |

[a] Aged <18 years.
[b] At the date of laboratory confirmation of second SARS-CoV-2 reinfection.
[c] At the date of SARS-CoV-2 laboratory confirmation of SARS-CoV-2 infection.
[d] Time duration of pandemic waves: Second wave: Jun. 2-Oct. 6, 2020; Third wave: Oct. 7, 2020-Jan. 31, 2021; Fourth wave: Feb. 1-Jul. 23, 2021; Fifth wave: Jul. 24-Dec. 31, 2021; Sixth wave: Jan.-Feb. 2022; Seventh (ongoing) wave: Jul. 1, 2022-.
[e] Vaccinated with one dose of BNT162b2, 49 days before symptom onset of second SARS-CoV-2 reinfection.

**eFigure 1. The Proportion of Primary SARS-CoV-2 Infections that were Reinfected in Children, Vojvodina, Serbia, March 6, 2020-July 31, 2022.[a]**

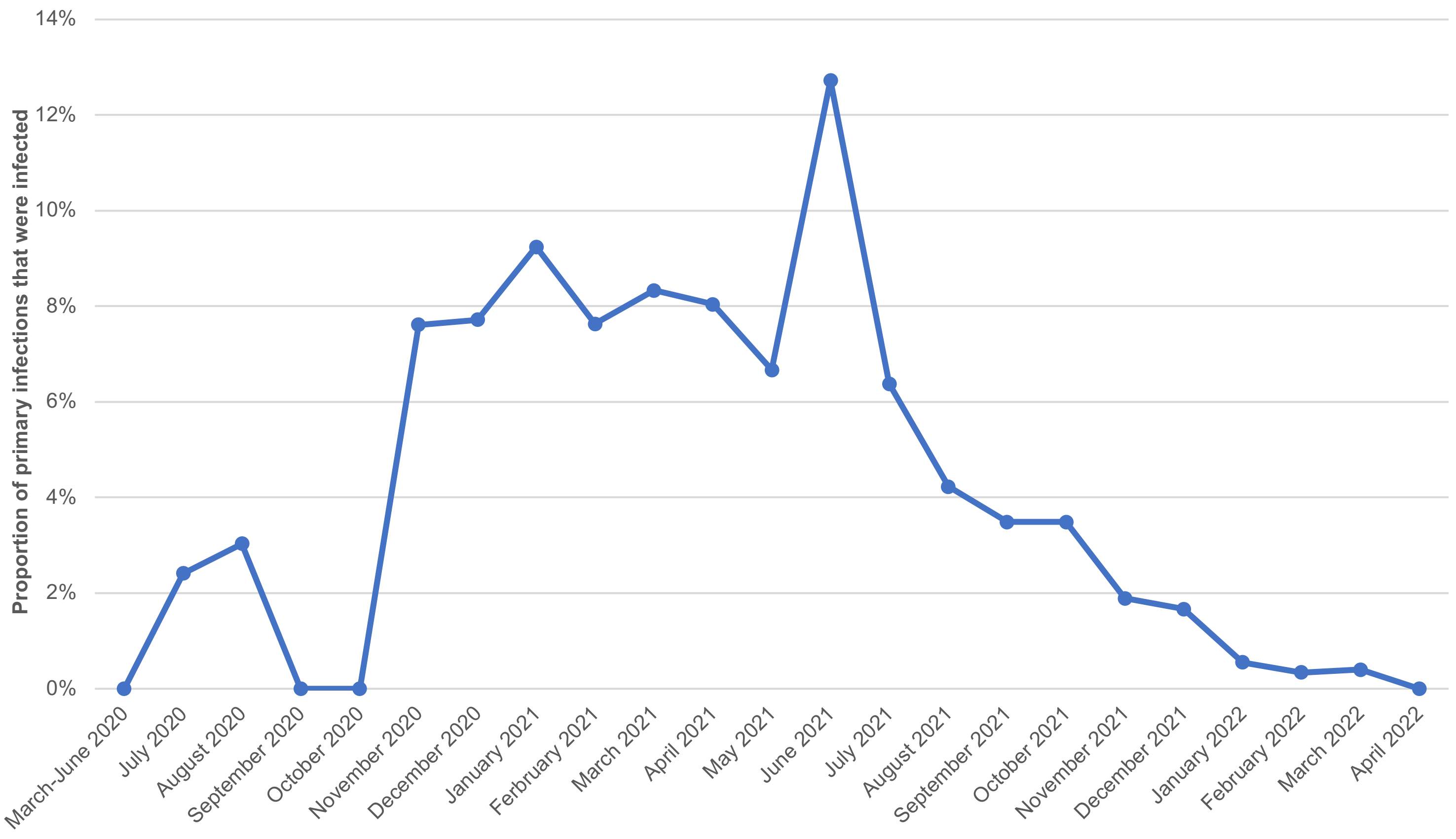


[a]Cochran–Armitage test was applied

**eFigure 2. Kaplan-Meier Curves showing the Cumulative Probability of Reinfection in the pediatric cohort (<18 years of age) According to (A) Severity of Primary Infection, (B) Age group and (C) Gender in Vojvodina, Serbia, March 6, 2020-July 31, 2022.**

A

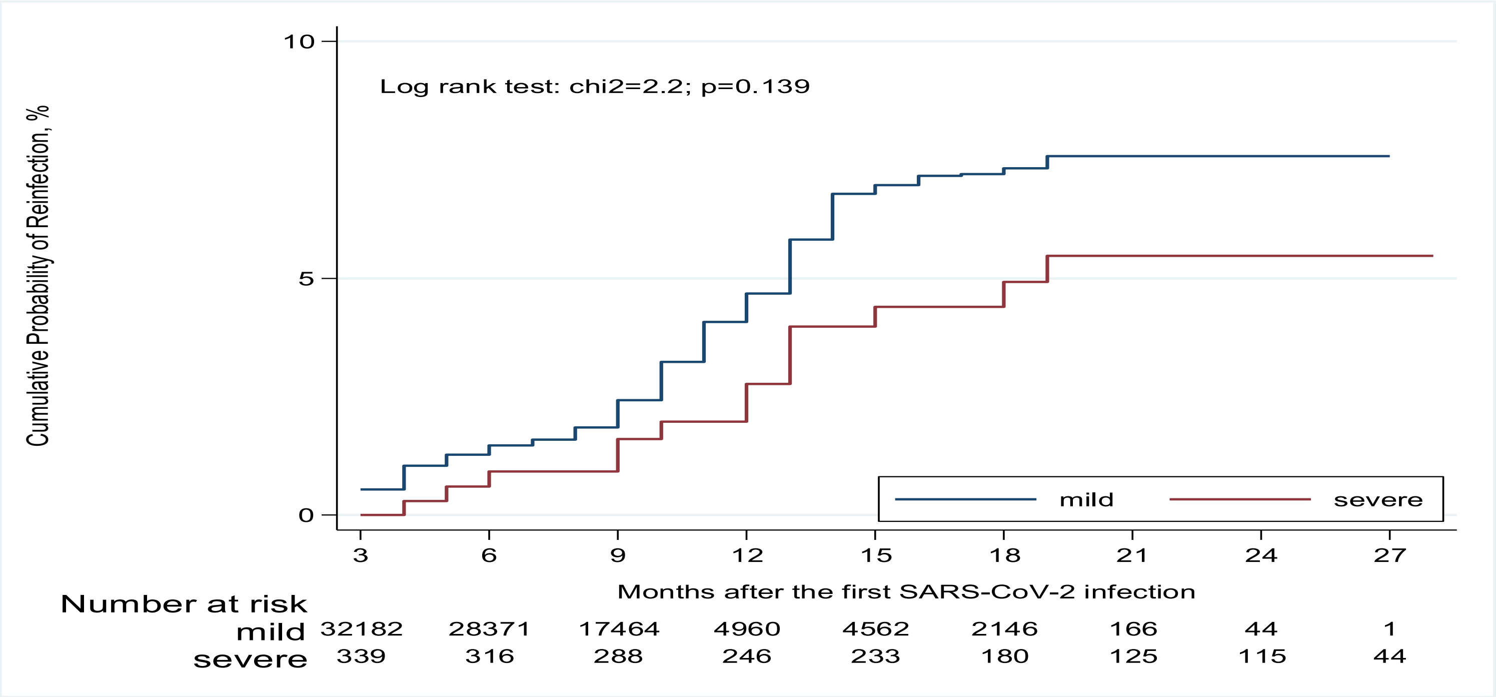

B

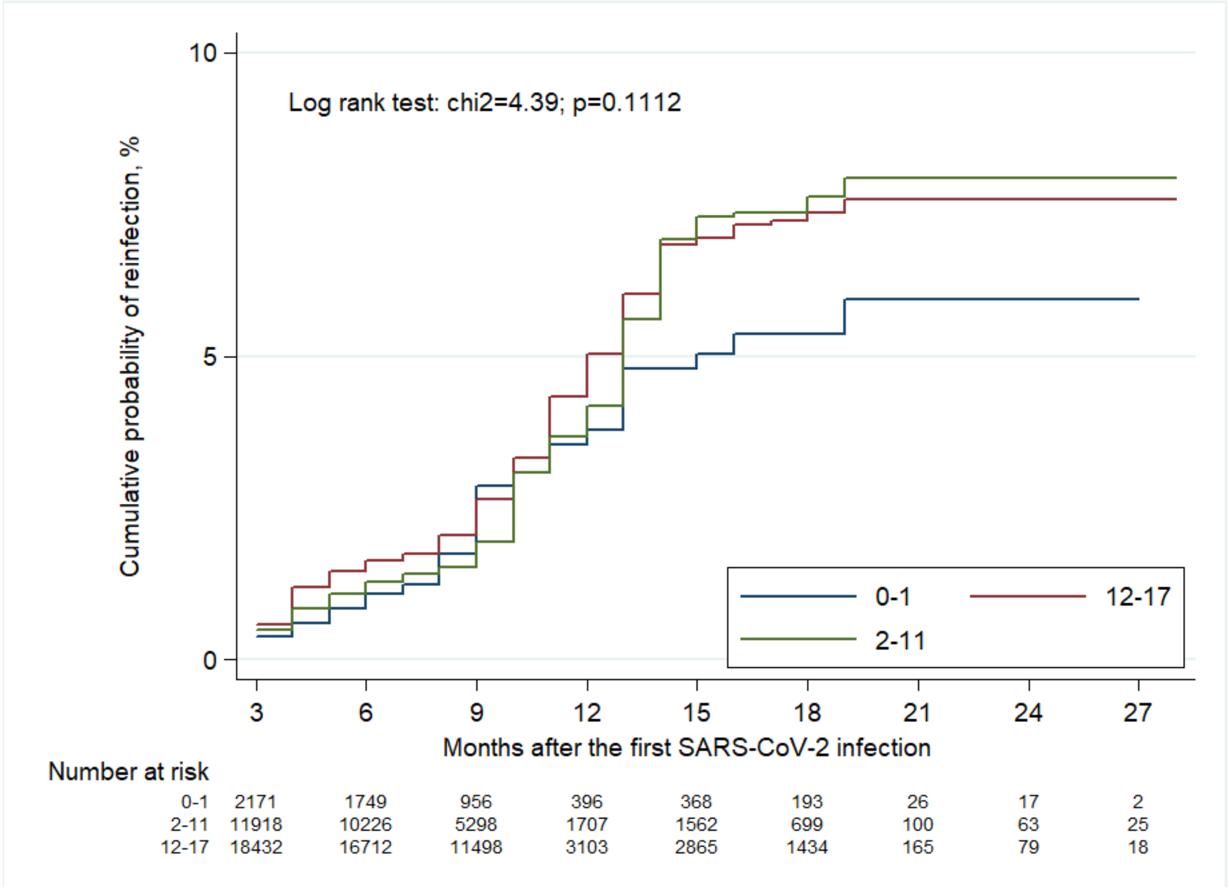
Log rank test: chi2=4.39; p=0.1112
Cumulative probability of reinfection, %
10
5
0
3
6
9
12
15
18
21
24
27
Months after the first SARS-CoV-2 infection
0-1
12-17
2-11
Number at risk
0-1 2171 1749 956 396 368 193 26 17 2
2-11 11918 10226 5298 1707 1562 699 100 63 25
12-17 18432 16712 11498 3103 2865 1434 165 79 18

C

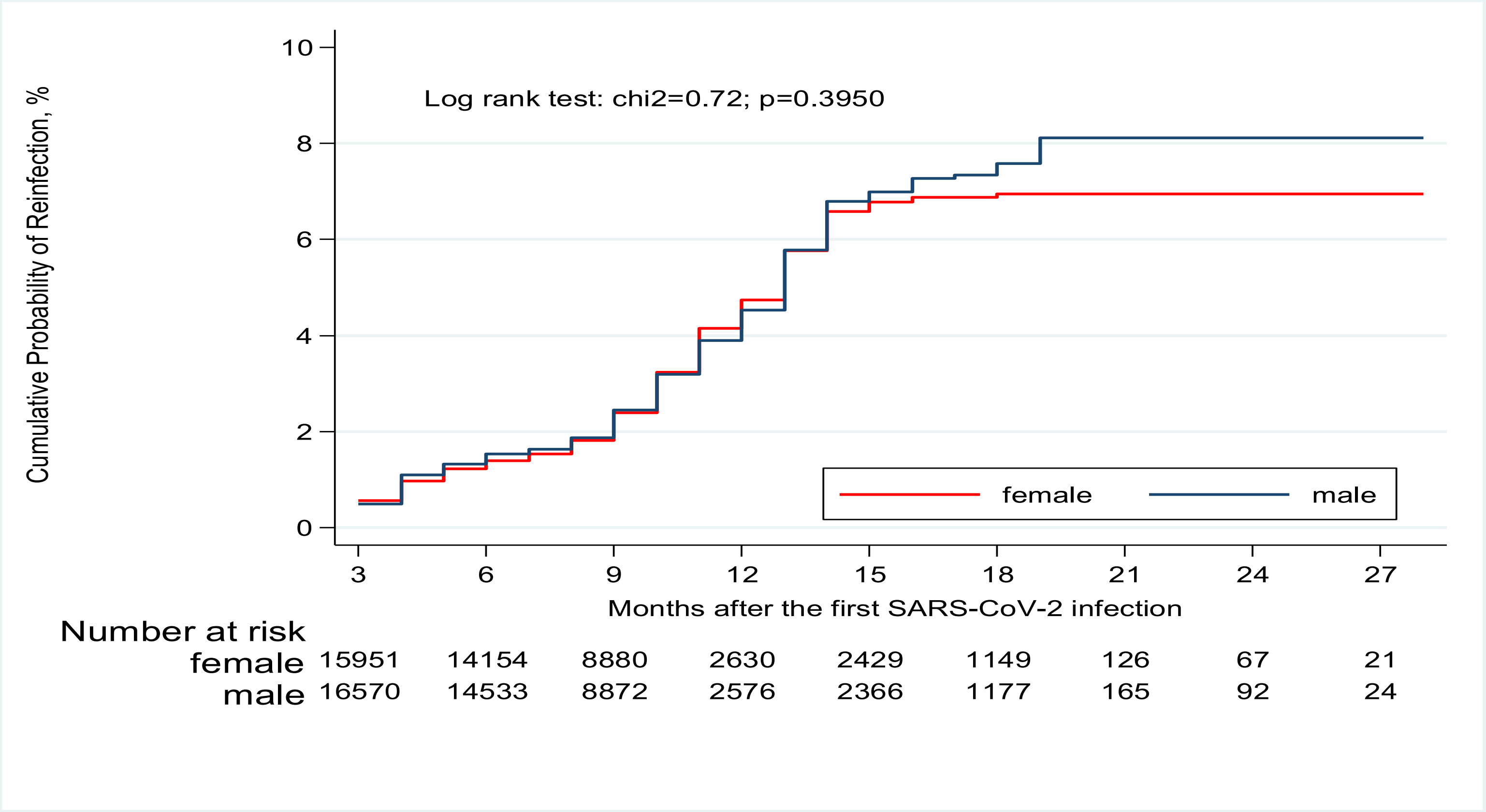
Cumulative Probability of Reinfection, %
10
8
6
4
2
0
Log rank test: chi2=0.72; p=0.3950
female
male
3
6
9
12
15
18
21
24
27
Months after the first SARS-CoV-2 infection
Number at risk
female 15951 14154 8880 2630 2429 1149 126 67 21
male 16570 14533 8872 2576 2366 1177 165 92 24

**eFigure 3. Severity of COVID-19 in Children and Adolescents (<18 years) without and with SARS-CoV-2 Reinfection (first and second episode), in Vojvodina, Serbia, March 6, 2020-July 31, 2022.**

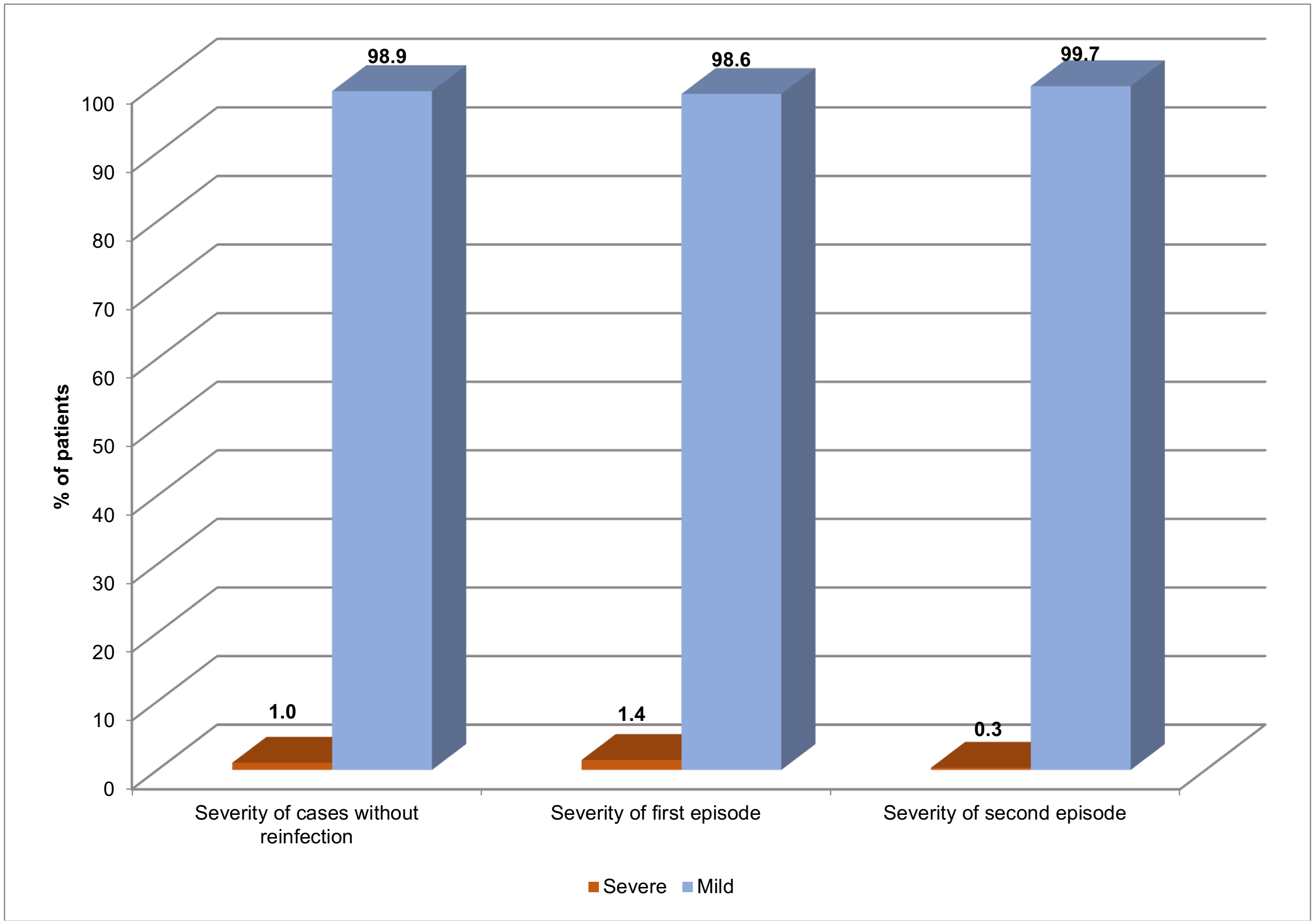

**eFigure 4. Kaplan-Meier Curves Showing the Cumulative Probability of Hospitalization (A) in the Pediatric Cohort and (B) for Patients with Reinfection in Vojvodina, Serbia, March 6, 2020-July 31, 2022.**

A

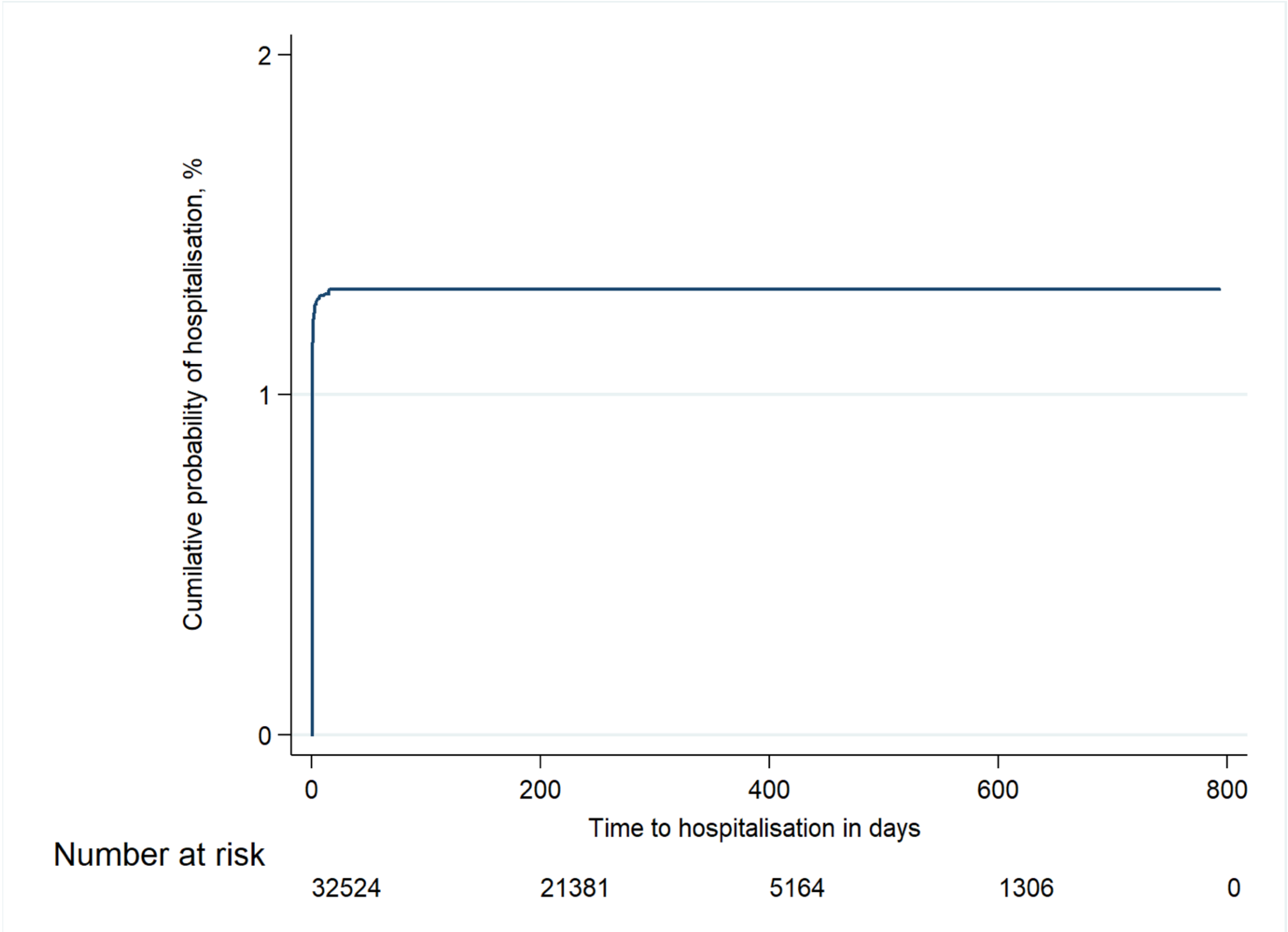

B

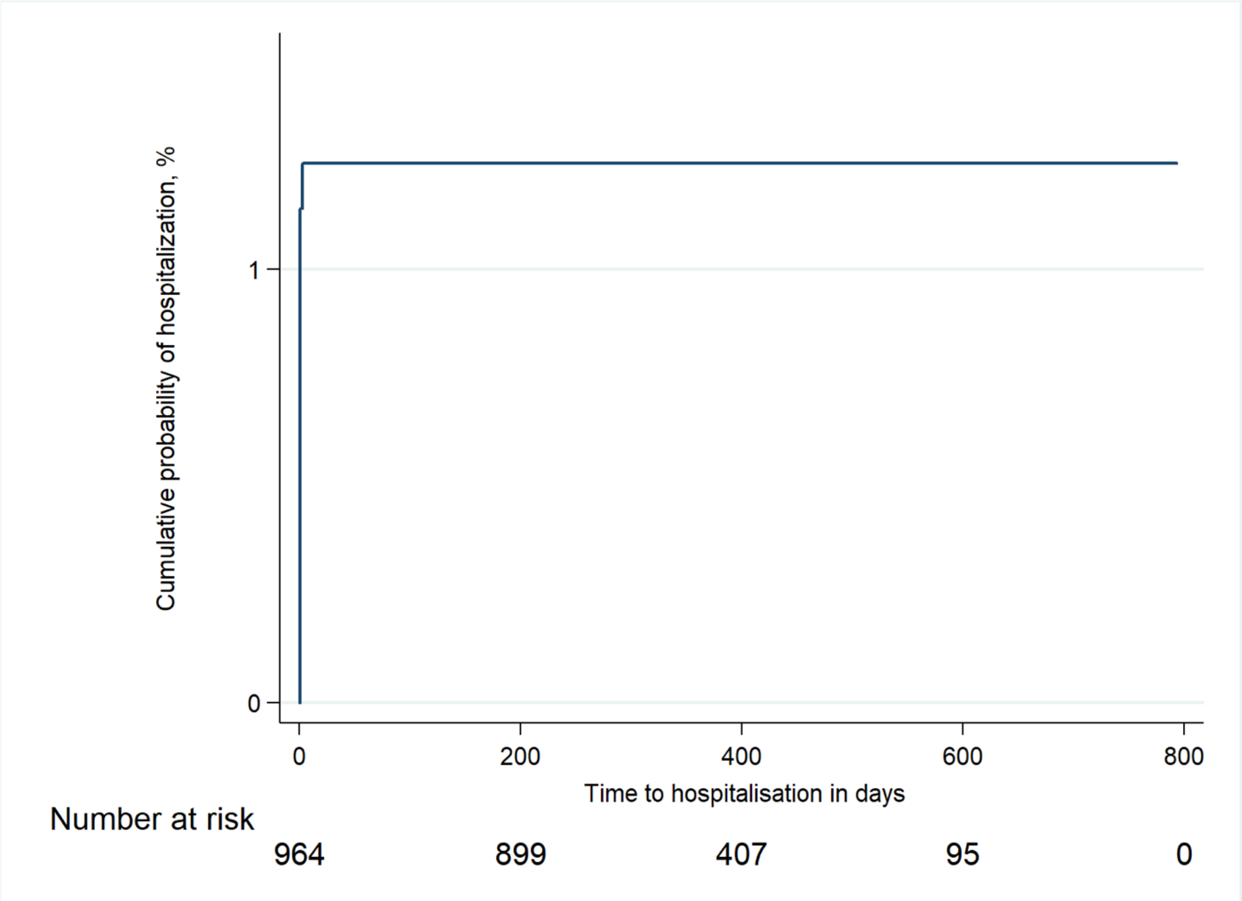
Cumulative probability of hospitalization, %
1
0
0
200
400
600
800
Time to hospitalisation in days
Number at risk
964
899
407
95
0
